# Circulating Markers of Angiogenesis and Endotheliopathy in COVID-19

**DOI:** 10.1101/2020.06.29.20140376

**Authors:** Matthew L. Meizlish, Alexander B. Pine, George Goshua, C-Hong Chang, Hanming Zhang, Jason Bishai, Parveen Bahel, Amisha Patel, Rana Gbyli, Jennifer Kwan, Christina Price, Charles S. Dela Cruz, Stephanie Halene, David van Dijk, John Hwa, Alfred I. Lee, Hyung J. Chun

## Abstract

Despite over 9.3 million infected and 479,000 deaths, the pathophysiological factors that determine the wide spectrum of clinical outcomes in COVID-19 remain inadequately defined. Importantly, patients with underlying cardiovascular disease have been found to have worse clinical outcomes,^1^ and autopsy findings of endotheliopathy as well as angiogenesis in COVID-19 have accumulated.^2,3^ Nonetheless, circulating vascular markers associated with disease severity and mortality have not been reliably established. To address this limitation and better understand COVID-19 pathogenesis, we report plasma profiling of factors related to the vascular system from a series of patients admitted to Yale-New Haven Hospital with confirmed diagnosis of COVID-19 via PCR, which demonstrate significant increase in markers of angiogenesis and endotheliopathy in patients hospitalized with COVID-19.

---

The protocol was approved by the Yale University institutional review board (IRB #2000027792). Blood was collected between April 13 and April 24, 2020 from critically ill patients (defined by admission to the intensive care unit at the time of blood draw, designated ‘ICU’, n = 40) and non-critically ill patients (‘non-ICU’, n = 9) with COVID-19 (**Figure 1A**). Blood from an additional 13 asymptomatic, non-hospitalized individuals were collected and served as controls (IRB #1401013259). Blood was collected in sodium citrate tubes and centrifuged at 4000 rpm for 20 minutes and the resulting plasma supernatant was used for further testing. The biomarker profiling analyses were conducted at Eve Technologies (Calgary, Alberta, Canada). Of the critically ill patients, 12 had died by the time of last follow-up on May 23, 2020, while 25 were discharged and 3 remained hospitalized. All of the non-ICU patients with COVID-19 had been discharged home, and none progressed to critical illness.

**Figure 1.**
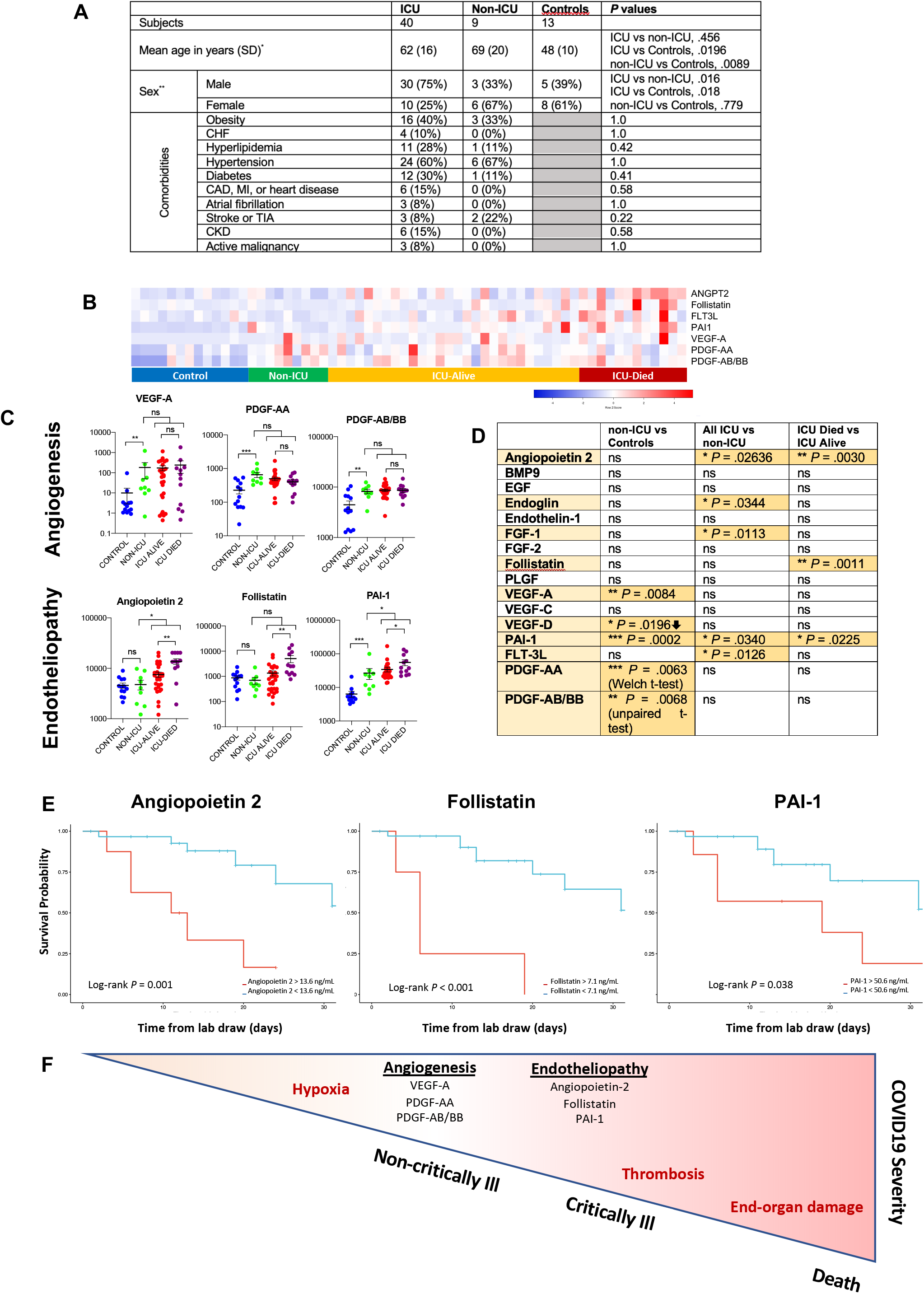
Analyses of vascular biomarkers across the spectrum of COVID-19 disease severity. (A) Subject demographics for ICU patients with COVID-19, non-ICU patients with COVID-19, and non-COVID-19, non-hospitalized controls. (B) Heatmap indicating relative protein levels detected in each subject (columns) for proteins (rows) that had statistically significant differences between groups. ICU patients are divided according to those who survived and those who died. (C) Comparisons of absolute plasma protein levels for select markers between control, non-ICU, and ICU groups, as well as ICU survivors vs. non-survivors; *\* P* < 0.05, *\*\* P* < 0.01, *\*\*\* P* < 0.001, ns: not significant. (D) Table of vascular markers tested for statistical significance between subject cohorts. (E) Kaplan Meier curves shown for angiopoietin-2, follistatin, and PAI-1, indicating that patients with elevated circulating levels of these proteins had a significantly higher likelihood of in-hospital mortality. (F) Schematic model of vascular processes in the progression of COVID-19 pathogenesis.

We carried out assessment of circulating markers related to vascular function, including markers of angiogenesis and endotheliopathy. A heatmap was generated using Heatmapper (heatmapper.ca) to represent the concentrations of circulating biomarkers (**Figure 1B**).^4^ We compared protein levels in 1) non-ICU vs. controls, 2) ICU vs. non-ICU, and 3) ICU non-survivors vs. ICU survivors (**Figures 1C**), and made three key observations relating vascular biomarkers with the severity of disease. First, multiple circulating pro-angiogenic factors, including VEGF-A, PDGF-AA, and PDGF-AB/BB, were significantly elevated in non-ICU COVID-19 patients compared to controls, which may be contributing to the recently described vascular remodeling processes in COVID-19 (**Figure 1C**).^3^ Second, in ICU COVID-19 patients, we found significantly increased levels of angiopoietin-2, FLT-3L, and PAI-1, which when present at high levels in circulation likely reflect ongoing endotheliopathy, and which support emerging data implicating endothelial involvement in critical illness (**Figure 1C)** (Goshua, et.al., *in press*).^2,3^ Other markers related to angiogenesis and endothelial function were also modestly changed or unchanged (**Figure 1D**). Third, we performed Kaplan-Meier survival analyses within the ICU population using unbiased cutpoints determined by maximally selected rank statistics.^5^ Markers of endotheliopathy segregated significantly with in-hospital mortality (angiopoietin-2, follistatin, PAI-1), suggesting that endotheliopathy may be a predictor of mortality in COVID-19 (**Figure 1E**).

Overall, we have identified a panel of circulating markers associated with angiogenesis and endotheliopathy, which increase at distinct stages of disease severity in COVID-19. We observe increased levels of angiogenic factors in hospitalized patients who are not critically ill, which may reflect hypoxemia that develops in many COVID-19 patients who require hospitalization. In patients with critical illness, we find circulating evidence of endothelial injury, which may drive other features of critical disease, including thrombosis and multi-organ failure (**Figure 1F**). Indeed, multiple biomarkers of endotheliopathy segregate with increased mortality. While a number of biomarkers, including D-dimer, troponin, and B-type natriuretic peptide, have been associated with survival in COVID-19, there is currently a paucity of vascular-specific biomarkers that can help prognosticate patients with COVID-19. This is a critical unmet need, given the emerging evidence for endothelial cell involvement in COVID-19 pathogenesis. Development of circulating markers that can detect specific aspects of COVID-19 pathogenesis may be critical to guide the use of novel therapeutic strategies, including those that may safeguard the vasculature, such as dipyridamole or inhibitors of the complement cascade. Further validation of our findings in larger patient cohorts, together with mechanistic studies to understand the causes of endothelial injury and its consequences for immune activation, vascular dysfunction, and thrombosis, promise to provide pivotal insights into COVID-19 pathogenesis and guide clinical management.

Supported in part by the National Heart Lung Blood Institute (HL142818) and gift donation from Jack Levin to the Benign Hematology Program at Yale.

## Data Availability

N/A

## References

1. Zhou F, Yu T, Du R, et al. Clinical course and risk factors for mortality of adult inpatients with COVID-19 in Wuhan, China: a retrospective cohort study. Lancet 2020;395:1054–62.

2. Varga Z, Flammer AJ, Steiger P, et al. Endothelial cell infection and endotheliitis in COVID-19. Lancet 2020;395:1417–8.

3. Ackermann M, Verleden SE, Kuehnel M, et al. Pulmonary Vascular Endothelialitis, Thrombosis, and Angiogenesis in Covid-19. N Engl J Med 2020.

4. Babicki S, Arndt D, Marcu A, et al. Heatmapper: web-enabled heat mapping for all. Nucleic Acids Res 2016;44:W147–53.

5. Hothorn T, Lausen B. On the exact distribution of maximally selected rank statistics. Computational Statistics & Data Analysis 2003;43:121–37.

